# Mental Health Trajectories of Occupational Physicians in the COVID-19 Pandemic: A Reciprocal Path Model of Burnout, Depression, and Anxiety

**DOI:** 10.1101/2025.05.17.25327758

**Authors:** João Silvestre Silva-Junior, Paulo Sergio Panse Silveira, Alberto Jose Niituma Ogata, Rodrigo Bornhausen Demarch, José Oliveira Siqueira

## Abstract

**Objective:** To propose and test psychometric models establishing direct, cross-lagged, and reciprocal longitudinal relationships between burnout, depression, and anxiety among occupational physicians during the COVID-19 pandemic.

**Methods:** This one-year prospective cohort study used repeated online questionnaires at three time points (2020–2022). A convenience sample of occupational physicians was recruited through professional networks and scientific associations. Data collection included a sociodemographic questionnaire, the Stanford Professional Fulfillment Index (PFI), and the Depression, Anxiety, and Stress Scale (DASS-21). Path analysis using Reciprocal Cross-Lagged Panel Models (RCLPM) assessed dynamic, bidirectional relationships among psychological variables over time, controlling for age, sex, and years of practice. Three models were specified, each centered on burnout, anxiety, or depression, to determine the best fit to the data.

**Results:** Of 540 participants, 124 completed all waves; 435 participated in wave 1, 220 in wave 2, and 265 in wave 3. Most participants were women, with similar age and experience across waves. Burnout declined from T1 to T2 and remained stable at T3. Depression and anxiety scores consistently decreased. All models revealed significant reciprocal cross-lagged effects, with feedback loops indicating mutual reinforcement. Anxiety emerged as particularly influential over time. Model fit indices confirmed model robustness, and stability indices suggested psychological stabilization over time.

**Conclusion:** Burnout, depression, and anxiety are reciprocally interrelated and evolve dynamically among occupational physicians. Burnout appears central in these interactions. Findings highlight the need for integrated strategies addressing both individual and organizational contributors to mental distress, especially during crisis contexts such as the COVID-19 pandemic.

## INTRODUCTION

Burnout is a syndrome associated with chronic exposure to occupational stressors^1^. It is recognized as an important problem that compromises the health of workers, particularly in the healthcare sector. In this group of workers, it has been associated with low-quality service provision^2^ and medical errors^3–5^.

In the United States of America (USA), annual losses of approximately 4.6 billion dollars are attributed to burnout, due to physician turnover—when professionals transition to other roles or leave medical practice—and reduced working hours, leading to vacant medical positions^6^. A systematic review analyzing data from 182 studies involving nearly 110,000 physicians found a burnout prevalence of 67%^7^. Despite the need to assess physicians’ well-being, multiple definitions of burnout exist, leading to the use of different assessment questionnaires^1^. Consequently, associations between burnout and factors such as sex, age, location, workload, medical specialty, and depressive symptoms pose challenges for comparability across published studies^7^.

Burnout and depressive disorders are often closely associated, with some studies suggesting that they may represent different manifestations of the same underlying problem^4, 5^. A review of 92 studies found that the association between burnout and depression remains conceptually uncertain, with some studies supporting the hypothesis that burnout is distinct from depression^5^. However, a systematic review and meta-analysis identified positive and statistically significant associations between burnout and depression, as well as between burnout and anxiety, linking burnout to the presence of depression and anxiety among healthcare professionals while still suggesting that they are distinct constructs^8^. In the context of Brazilian workers, the relationship between burnout and depression remains poorly understood.

In 2022, there were more than 562,000 registered physicians in Brazil. Among them, over 20,000 specialized in Occupational Medicine^9^. One of the primary responsibilities of these professionals is managing health promotion and disease prevention programs that employers are required to implement within their organizations. According to a review by Eisch et al., common stressors in occupational medicine practice include organizational deficiencies, economic uncertainties, intense professional responsibilities, and the need to maintain high standards in poorly structured work environments. These factors can negatively impact the mental and physical health of these professionals, affecting the effectiveness of occupational health services^10^.

These challenges were exacerbated during the COVID-19 pandemic. A comprehensive global review encompassing 250 studies revealed alarming pooled prevalence rates of anxiety (43.6%), depression (37.1%), and burnout (43.6%) among healthcare workers in the pandemic context^11^. A Brazilian survey of healthcare professionals found that the perception of negative psychosocial aspects at work, such as high task demands with low decision latitude and low coworker support, increased the likelihood of mental distress among workers in the first year of the pandemic^1^].

In this context, the objective of this study is to propose and test psychometric models that establish direct, cross-lagged, and reciprocal longitudinal relationships between burnout, depression, and anxiety among occupational physicians during the COVID-19 pandemic.

## METHODS

This study can be defined as a short-term prospective cohort, as it is an observational study in which questionnaires were repeatedly administered to assess changes in burnout, depressive symptoms, and anxiety scores. Data collection was conducted in three waves: T1 – from October 2020 to January 2021; T2 – from May to September 2021; and T3 – from November 2021 to January 2022.

### Participants

The study used a convenience sample, with the invitation disseminated through multiple social networks of professionals involved in human resources training in the field, as well as through scientific associations representing the specialty of Occupational Medicine at both national and regional levels. These associations also forwarded the invitation to their members via email.

### Data collection

At the time of entry into the research, the participants responded to online questionnaires, made available through a link on the Google Forms, regarding:

- Sociodemographic characterization: date of birth, sex, length of professional practice in the Occupational Medicine field;
- Stanford Professional Fulfillment Index (PFI) Brazilian Portuguese version of the questionnaire with 16 sentences and response options on a five-point Likert scale that assesses two dimensions (professional fulfillment and burnout)^13,14^;
- Information on depression and anxiety was obtained through the completion of the Brazilian version of the short form of the Depression, Anxiety, and Stress Scale (DASS-21), which includes seven questions for each symptom group referring to the week prior to the survey^15, 16^.

### Longitudinal approach

Participants who entered wave 1 were invited to participate in wave 2 after 180 days and in wave 3 after 365 days after wave 1. Participants who missed the first wave could enter Phase 2 and were invited to be part of wave 3 after 180 days. In all waves, participants completed the PFI and DASS-21 questionnaires.

### Statistical analysis

A path analysis model was chosen to assess the reciprocal relationships between burnout, anxiety, and depression simultaneously, as well as their effects over the three waves. Path analysis is a specialized subset of structural equation modeling (SEM) used in statistics to describe the directed dependencies among a set of overt variables. This technique is particularly useful in cases where a researcher is interested in examining hypotheses about the relationships among overt variables, particularly causal models. Path analysis allows for the estimation of the strength and significance of causal connections between variables. It is commonly used in social sciences, economics, and other fields to understand complex relationships where multiple variables influence each other directly and indirectly. Unlike traditional regression models, path analysis can include multiple dependent variables and allows for the examination of interrelations among all variables in the model.

A nonrecursive SEM model is a type of statistical model used in social sciences, psychology, and other fields. Unlike recursive models, nonrecursive models allow for feedback loops and bidirectional relationships between variables. This means that variables in a nonrecursive SEM can both influence and be influenced by other variables in the model. These models are more complex and can better capture the dynamic and interdependent nature of real-world phenomena. They are used when the assumption of unidirectional causality (present in recursive models) does not hold, allowing for a more nuanced understanding of the relationships between variables.

The Reciprocal Cross-Lagged Panel Model (RCLPM) model is a statistical technique used in longitudinal studies to analyze the dynamic, including bidirectional relationships between variables. It involves measuring the same variables at multiple time points and examining how each variable at one time point influences the other variables at subsequent time points, while also accounting for previous influences. This model helps to understand the interplay and reciprocal influence between variables, revealing how they might impact each other over time in a cyclic or ongoing manner.

In the current study, burnout, anxiety, and depression may reinforce each other, which is the nonrecursive component that implements feedback loops, taking into account three time points that influence subsequent variables over time. It is important to note that this kind of modeling does not impose a transversal causation order (e.g., burnout may cause or be a consequence of anxiety and/or depression); however, it preserves longitudinal reinforcements (e.g., burnout leading to more burnout or increasing depression and anxiety from one moment to the subsequent). IBM SPSS AMOS 24^17^ was chosen because it is among the rare software packages capable of processing non-recursive models. This situation presents a common challenge in empirical research, particularly in the context of Structural Equation Modeling (SEM), where missing data can significantly impact the validity of the results.

### Ethics aspect

The study was approved by the Human Research Ethics Committee of São Camilo University Center (Opinion 4,077,484/2020), with registration on the Brazil Platform (CAAE 31400620.0.0000.0062). All eligible participants gave their consent to participate after reading and accepting the Informed Consent Form.

## RESULTS

In the statistical analysis of a dataset comprising a total of 540 cases, it was identified that only 124 cases (22.96%) are complete with no missing values. Additionally, there are 4053 valid observations, which constitute 62.55% of the data.

The first wave had the highest number of participants (435). Among them, 127 took part in both the second and third waves, 58 participated only in the second wave, and 57 participated only in the third wave. In other words, 193 participants were exclusive to the first wave. Regarding the 220 participants in the second wave, 11 also participated in the third wave. Among the 265 participants in the third wave, 70 joined the study only at this stage (Figure 1).

**Figure 1.**
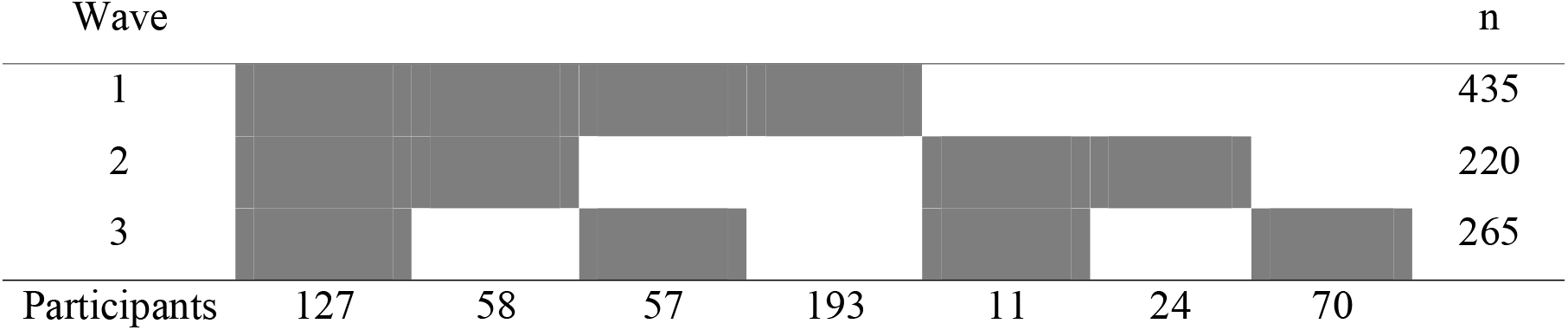
Distribution of participants according to participation in each wave of the study, 2020-2022, Brazil (n=540)

Women were the majority in all waves. Age distribution was similar across groups, as was the length of time working in the specialty. The burnout score dropped considerably between the first and second waves, remaining stable in the third. Depression scores showed a sustained decline, as did anxiety scores (Table 1).

**Table 1.** Description of the participants according to survey waves, 2020-2022, Brazil (n=540)

| Variable | Wave 1<br>(n=435) | Wave 2<br>(n=220) | Wave 3<br>(n=265) |
| --- | --- | --- | --- |
| **Female** | 57.9% | 57.7% | 56.2% |
| **(Mean, standard deviation)** |  |  |  |
| **Age** | 49.6 (14.0) | 47.1 (14.7) | 49.6 (13.5) |
| **Time of employment** | 13.7 (9.7) | 12.7 (9.1) | 14.5 (10.1) |
| **Burnout score** | 12.4 (8.7) | 10.3 (7.4) | 10.7 (7.7) |
| **Depression score** | 5.3 (5.1) | 4.9 (4.6) | 4.1 (4.2) |
| **Anxiety score** | 3.4 (4.0) | 2.8 (3.4) | 2.4 (3.1) |

The following section presents three Panel Models of Reciprocal Correlation with Cross-Lagged Effects, in which burnout or depression or anxiety are positioned on the central axis and tracked over time. In each of these models, the other variables are incorporated as feedback loops, exerting influence on their subsequent time points.

In Figure 2 burnout occupies the central axis that follows over time. Depression and anxiety are implemented as feedback loops and influence their subsequent moments. Burnout influences the three variables in the subsequent moments. Age, sex and time of employment are control variables (RMSEA = 0.05 [0.04, 0.07], pclose = 0.27, NFI = 0.96, Stability indices: wave1 = 0.6, wave2 = 0.16, wave3 = 0.25).

**Figure 2.**
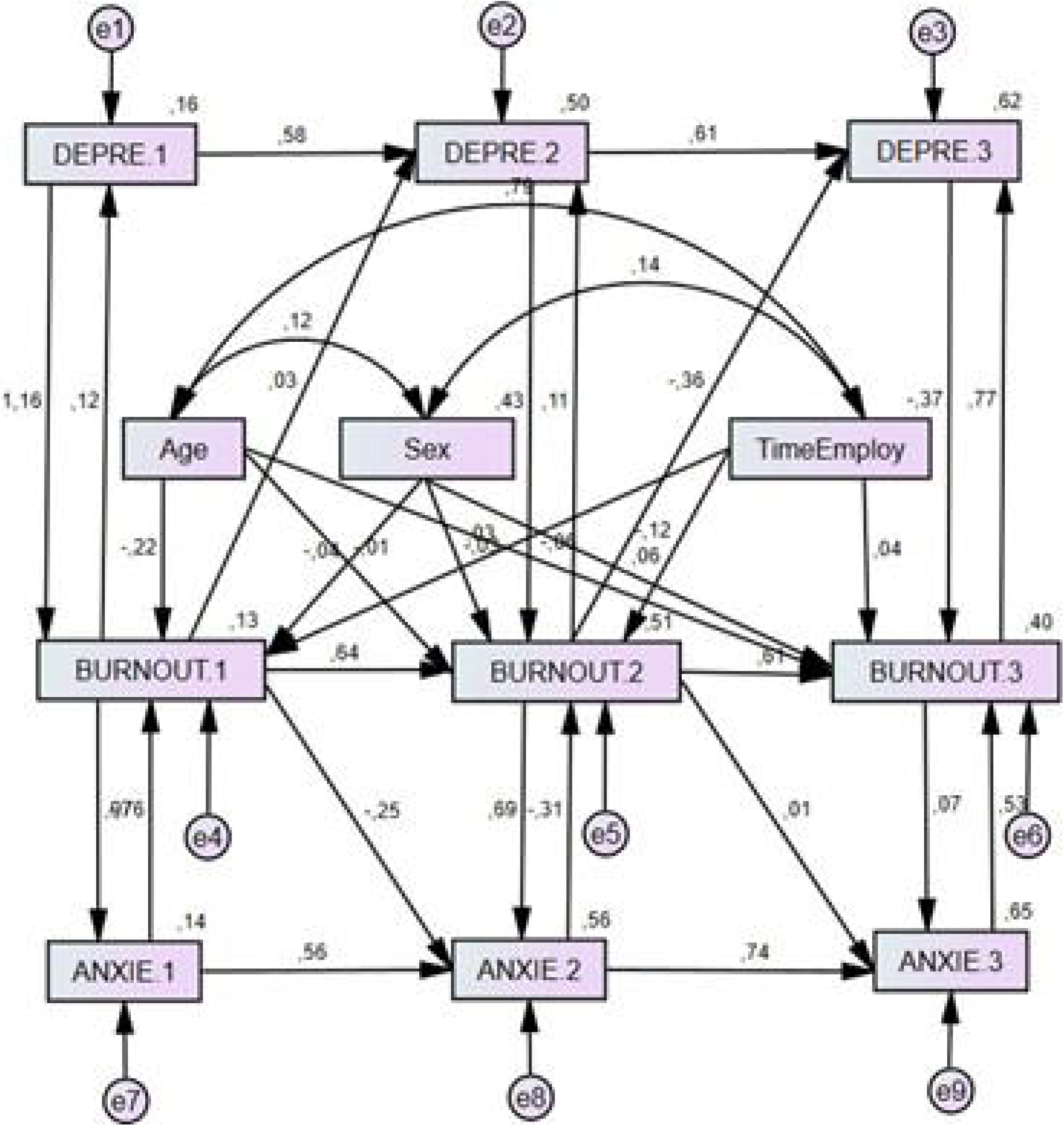
Reciprocal Cross-Lagged Panel Correlation model in which burnout occupies the central axis that follows over time.

In Figure 3 depression occupies the central axis that follows over time. Anxiety and burnout are implemented as feedback loops and influence their subsequent moments. Depression influences the three variables in the subsequent moments. Age, sex and time of employment are control variables (RMSEA = 0.07 [0.06, 0.08], pclose = 0.006, NFI = 0.95, Stability indices: wave1 = 2473.44, wave2 = 0.23, wave3 = 0.37).

**Figure 3.**
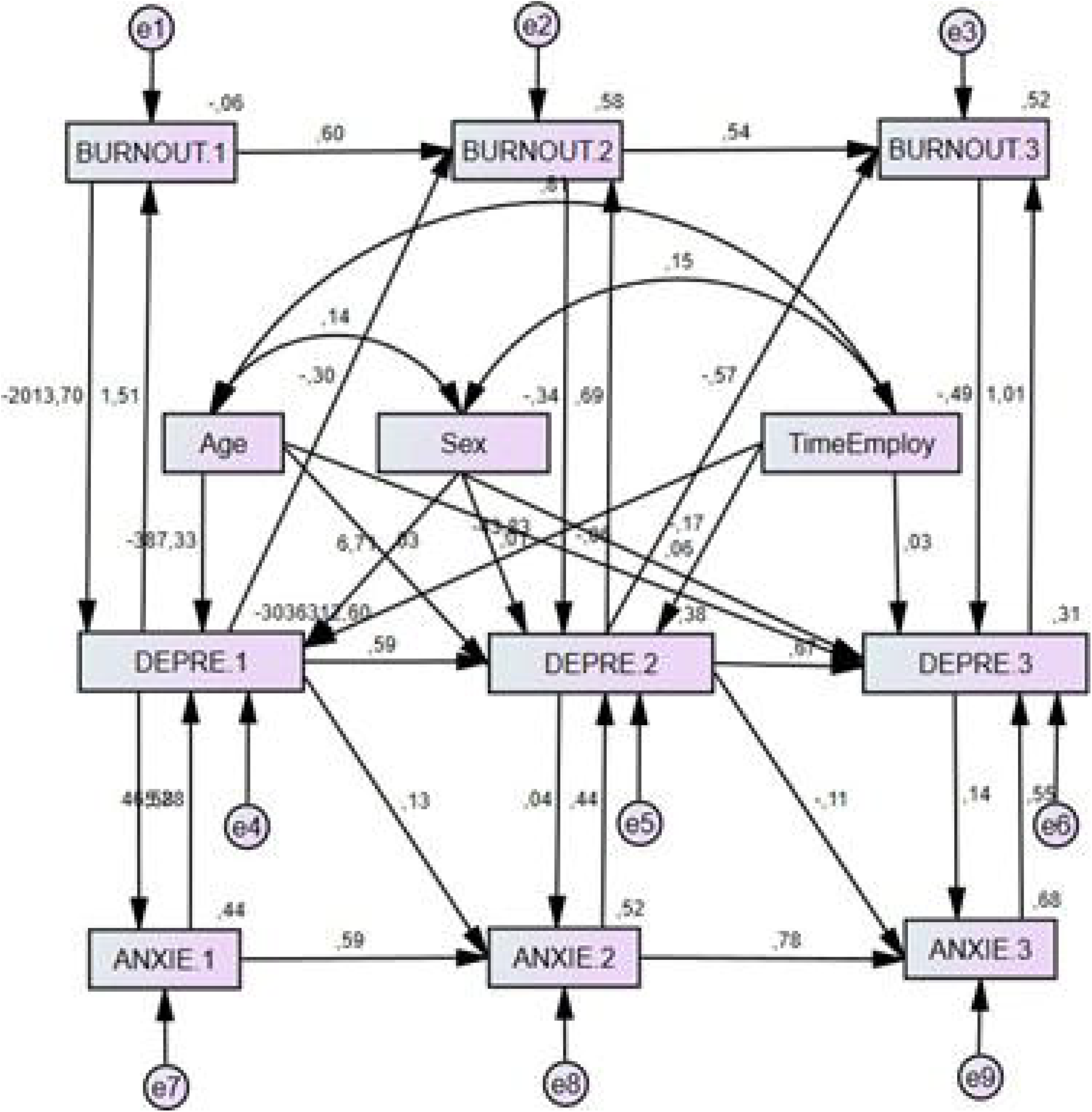
Reciprocal Cross-Lagged Panel Correlation model in which depression occupies the central axis that follows over time.

In Figure 4 anxiety occupies the central axis that follows over time. Burnout and depression are implemented as feedback loops and influence their subsequent moments. Anxiety influences the three variables in the subsequent moments. Age, sex and time of employment are control variables (RMSEA = 0.06 [0.05, 0.08], pclose = 0.056, NFI = 0.96, Stability indices: wave1 = 18.54, wave2 = 0.43, wave3 = 0.86).

**Figure 4.**
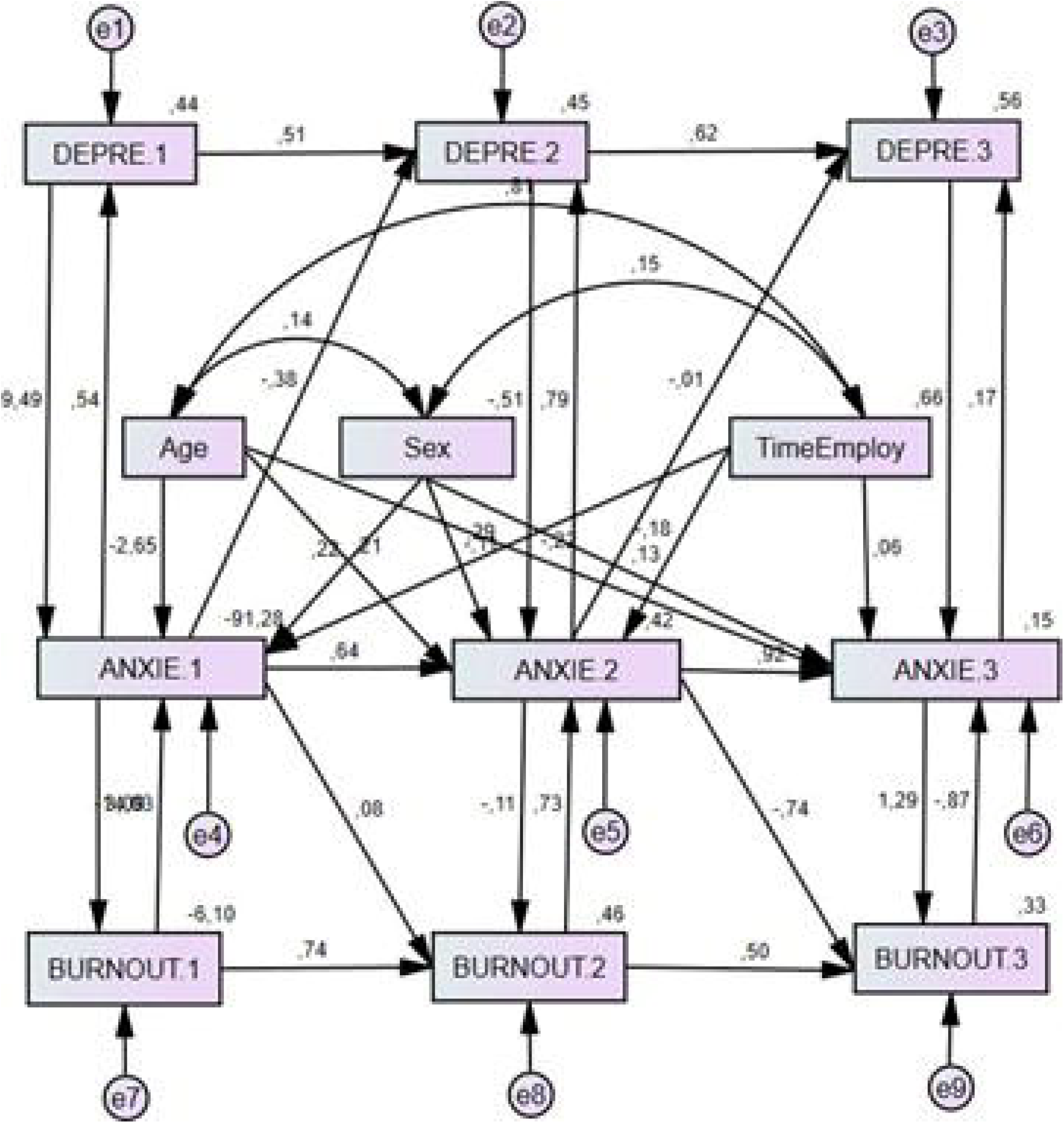
Reciprocal Cross-Lagged Panel Correlation model in which anxiety occupies the central axis that follows over time.

The analysis reveals that all three models exhibit contemporary reciprocal relationships, characterized by cross-lagged feedback effects. The stability indices indicate a convergence at infinity, suggesting that the variables tend to stabilize over time. Furthermore, the results demonstrate that each variable influences the other both contemporaneously and across time. Age, gender, and length of employment are included as control variables to account for potential confounding effects.

## DISCUSSION

This study applies path analysis with reciprocal influences among burnout, depression, and anxiety in a cross-lagged panel to explore their relationship over time. In the literature, some applications are cross-sectional, such as examining the relationship between attitude toward mathematics and achievement using path analysis^18^ or assessing mutual feedback between happiness and life satisfaction through non-recursive confirmatory analysis^19^. A longitudinal approach appears in a study on depressive symptoms and alcohol consumption at two time points, using path analysis with feedback between them only at the final stage^20^. Other longitudinal studies involve methodological approaches in medical research that apply structural equation models but do not incorporate reciprocity between latent variables^21–23^. Combining contemporaneous feedback with longitudinal influences is particularly relevant, as seen in a study on postpartum depression and marital conflict^24^.

The central issue is determining whether anxiety, depression, and burnout are distinct constructs or aspects of the same construct and, if they are distinct, what their relationship is. Given that they are, it is necessary to examine whether there are contemporaneous and lagged effects. Evidence supports the first hypothesis, namely that they are three distinct but interrelated constructs^8, 25–27^. The present study contributes to the second question.

Path analysis was chosen because it is a genuinely multivariate model with a global test that generates an *omnibus p*-value. It differs from the regressions in a univariate general linear model because, instead of treating multiple independent variables as potential explanations for a single dependent variable (outcome), it allows for the analysis of relationships between outcomes. This analytical framework creates a network of effects: direct effects (when there are reciprocal, contemporaneous relationships, e.g., burnout affects anxiety, and anxiety affects burnout), and longitudinal relationships defined either by the passage of time (when a manifest variable influences another of the same nature, e.g., burnout at one time point affecting burnout later) or by cross-lagged effects (when variables of one type influence variables of another type, e.g., whether anxiety or burnout influences the subsequent state of burnout or anxiety, respectively). Additionally, this modeling approach generates numerous indirect effects, as multiple pathways between any two variables emerge through intermediaries. This approach enables the evaluation of the significance of each possible pathway within the overall model rather than focusing solely on the statistical significance of pairwise relationships. These coe□icients are represented alongside the arrows in Figures 2, 3, and 4.

When processed in the AMOS environment, this modeling approach is computationally intensive, requiring multiple iterations. Reciprocal relationships must converge until they reach stability in their mutual influences, eliminating the traditional distinction between cause and effect (i.e., the contemporary concept of causality no longer applies). However, since the model also includes direct and cross-lagged longitudinal relationships, the computation must determine valid parameter estimates to link the results from the three waves of questionnaires administered to participants. Among the three models tested, acceptability and stability conditions are satisfied only when burnout occupies the central axis of the path analysis diagram, i.e., the model is not rejected based on the *p*-value of the RMSEA, with pclose *>* 0.05, and the stability indices remain between -1 and +1 across all waves (Fig. 2).

Similarly to the results found in this research with a group of doctors, other longitudinal studies have demonstrated a bidirectional interaction between burnout and depressive symptoms. Bianchi et al.^5^ review highlights that the circular nature of the relationship between burnout and depression has been reported in multiple longitudinal studies, reinforcing the hypothesis of an interdependence between these phenomena. Ahola and Hakanen^28^ identified a reciprocal relationship between burnout and depression among Finnish dentists, suggesting that occupational stress predisposes individuals to depression through burnout, while burnout can be exacerbated both directly and indirectly by depression. McKnight and Glass^29^ replicated this finding among nurses in the United States, demonstrating that approximately 20% of the shared variance between burnout and depression can be attributed to their joint development, without a definitive temporal sequence. Toker and Biron^30^ corroborated this reciprocal relationship in a longitudinal study with Israeli workers, where increases in depressive symptoms predicted subsequent increases in burnout and vice versa. Koutsimani et al. ^8^ argue in their review that burnout is more strongly related to the occupational context, whereas depression is a generalized condition. That is, the process of mental health degeneration from depression may be triggered by general stressors and be related to burnout when occupational stressors are present.

As for the results of the reciprocal relationship between burnout and anxiety found in this study, the literature also suggests a significant and complex connection between the two, with evidence indicating that burnout and anxiety mutually influence each other over time, often exacerbating mental health issues and contributing to a cycle of persistent stress and emotional strain. Koutsimani et al. ^8^ indicate that burnout and anxiety are distinct constructs. According to this review, several cross-sectional studies provide evidence that occupational stress can increase the likelihood of developing anxiety symptoms, particularly in the presence of high job demands and excessive commitment. In the baseline of this research, participants with excessive work commitment showed a higher likelihood of experiencing anxiety symptoms^31^. Studies further explore this relationship by considering the three components of burnout: emotional exhaustion, cynicism, and low professional e□icacy. Ding et al.^32^ found that the first two were positively associated with anxiety symptoms, whereas higher professional e□icacy was negatively related. Moreover, Turnipseed^33^ suggests that the interaction between working conditions and individual characteristics can foster states of anxiety and, by extension, contribute to the development of burnout.

In this study, the tool used to measure overall burnout was the Stanford Professional Fulfillment Index (PFI), which evaluates the dimensions of work exhaustion and interpersonal disengagement. The Maslach Burnout Inventory (MBI) is the most commonly used questionnaire in other studies, but it has been the subject of criticism in the literature. Bianchi et al.^5^ review questions the validity of the MBI, arguing that its items may not capture a distinct phenomenon but rather represent a conceptual overlap with depression and anxiety in traditional psychiatric diagnoses. However, numerous studies utilizing the MBI have shown that burnout, depression, and anxiety are distinct phenomena, as also observed in this study with Brazilian workers using the PFI.

### Limitations

This study has several limitations. First, the sample was limited to Brazilian physicians, which may restrict the generalizability of the findings to other professions or populations. The COVID-19 pandemic context may have also uniquely influenced participants’ experiences, potentially exaggerating burnout, anxiety, and depression symptoms. Additionally, the use of self-reported measures introduces response bias, as participants may underreport or overreport symptoms. While the longitudinal design included multiple waves of data, a longer follow-up period could have provided deeper insights into the long-term evolution of these conditions. Lastly, although key demographic variables were controlled for, other potential confounders, such as social support and work environment, were not considered, which may have impacted the observed relationships. Future studies should address these limitations by including a broader range of variables and extending the follow-up period.

## CONCLUSION

In conclusion, this study contributes to the growing body of research on the complex interrelations between burnout, depression, and anxiety among occupational physicians during the COVID-19 pandemic. By applying path analysis with reciprocal and cross-lagged effects, we provide a more nuanced understanding of how these constructs interact over time. Our findings highlight burnout as a pivotal and initiating factor in this triad, often preceding and amplifying symptoms of depression and anxiety.

These results strongly support the need for integrated interventions that place burnout prevention and management at the core of broader mental health strategies. Addressing both occupational and psychosocial stressors—through coordinated organizational and individual-level actions—is essential.

Future research should further investigate the mechanisms underlying these relationships, particularly workplace-specific determinants, and adopt longitudinal designs to capture their long-term dynamics. Worker health initiatives must include systematic burnout screening, structural interventions to reduce chronic job stress, and sustained mental health support—especially for high-risk professional groups. By centering burnout in occupational mental health strategies, we can foster more effective and lasting interventions.

## Data Availability

All data produced in the present study are available upon reasonable request to the authors

## Notes

### Competing Interest Statement

The authors have declared no competing interest.

### Funding Statement

This study was funded by CGP Brasil.

### Author Declarations

Human Research Ethics Committee of Sao Camilo University Center (Opinion 4,077,484/2020), with registration on the Brazil Platform (CAAE 31400620.0.0000.0062), gave ethical approval for this work

